# Sex hormone-binding globulin, testosterone and type 2 diabetes risk in middle-aged African women: exploring the impact of HIV and menopause

**DOI:** 10.1101/2024.12.25.24319619

**Authors:** Julia H. Goedecke, Clement Nyuyki Kufe, Maphoko Masemola, Mamosilo Lichaba, Ikanyeng D. Seipone, Amy E Mendham, Hylton Gibson, James Hawley, David M. Selva, Itai Magodoro, Andre Pascal Kengne, Tinashe Chikowore, Nigel J. Crowther, Shane A Norris, Fredrik Karpe, Tommy Olsson, Karl-Heinz Storbeck, Lisa K. Micklesfield

## Abstract

**Objectives:** Sex hormone-binding globulin (SHBG) and testosterone are differentially associated with type 2 diabetes (T2D) risk. We investigated whether these associations differ by HIV and menopausal status in Black South African women living with (WLWH) and without HIV (WLWOH).

**Design:** Cross-sectional observational.

**Methods:** Eighty one premenopausal (57 WLWOH, 24 WLWH) and 280 postmenopausal (236 WLWOH, 44 WLWH) women from the Middle-Aged Soweto Cohort (MASC) completed the following measures: circulating SHBG and sex hormones, body composition (dual energy x-ray absorptiometry), oral glucose tolerance test to estimate insulin sensitivity (Matsuda index), secretion (insulinogenic index, IGI) and clearance, and beta-cell function (disposition index, DI). Dysglycaemia was defined as either impaired fasting or postprandial glucose or T2D.

**Results:** SHBG was higher and total and free testosterone were lower in postmenopausal WLWH than WLWOH (all p≤0.023). Irrespective of HIV serostatus, SHBG was positively associated with Matsuda index, insulin clearance and DI and inversely with HOMA-IR (all p<0.011). The association between SHBG and Matsuda index was stronger in premenopausal than postmenopausal women (p=0.043 for interaction). Free testosterone (and not total testosterone) was only negatively associated with basal insulin clearance (p=0.021), and positively associated with HOMA-IR in premenopausal and not post-menopausal women (p=0.015 for interaction).

**Conclusions:** We show for the first time that midlife African WLWH have higher SHBG and lower total and free testosterone than WLWOH, which corresponded to their higher beta-cell function, suggesting a putative protective effect of SHBG on T2D risk in WLWH.

**Significance statement:** This study in midlife Black African women suggest that higher sex hormone binding protein (SHBG) and lower free testosterone in women living with HIV (WLWH) may be associated with reduced risk of type 2 diabetes (T2D) compared to women living without HIV. Further, this study provides a putative mechanism underlying the lower prevalence of T2D in WLWH and obesity compared to women living with obesity but without HIV. However, longitudinal studies are required to understand the clinical implications of these findings.

## Introduction

Type 2 diabetes (T2D) has reached epidemic proportions globally and is associated with obesity and ageing^1^. Sub-Saharan Africa (SSA) is the region with the highest projected relative increase in T2D (129% by 2045)^1^, with South Africa (SA)^2^ having the highest number of people living with T2D in SSA^3^. Midlife SA women are disproportionately affected by T2D^4^, with putative drivers being HIV, the menopausal transition, and the high prevalence of obesity^5^ ^6^. Hormonal changes emerge as a key factor connecting these health challenges, with studies suggesting that sex hormones play a critical role in the development and progression of T2D^7–11^.

The menopausal transition is characterised by an increase in the androgen to E2 ratio, which reflects the substantial decline in oestrogen (E2) production and a slower rate of decline in androgen biosynthesis^9^. Higher testosterone in women has been associated with increased risk for T2D^7, 11–13^. However, there is evidence to suggest that the association between testosterone and T2D risk may be driven (directly or indirectly) by the effects of sex hormone binding globulin (SHBG)^7, 14^. Indeed, recent Mendelian randomisation studies showed an independent association between SHBG and T2D^7, 8, 15^.

Given the impact of SHBG and androgens on the risk of T2D, it is important to investigate their potential independent roles in the prevalence of dysglycaemia among two high-risk groups; women living with HIV (WLWH) and postmenopausal women. On average, SHBG decreases over the menopausal period^16^, with SHBG being significantly lower in postmenopausal compared to premenopausal women^17^. Results from the Women’s Interagency HIV study (WIHS) have shown that WLWH have lower E2 and total testosterone, but higher SHBG compared to women living without HIV (WLWOH)^18–21^. However, this study did not explore the role of SHBG and androgens on insulin dynamics (i.e. insulin sensitivity, insulin response and beta-cell function), which is integral to the development of T2D^22^. We recently showed that SHBG was associated with incident T2D, insulin sensitivity and beta-cell function in a cohort of middle-aged Black SA men without HIV^23^. This may be because the already elevated SHBG levels in men living with HIV (MLWH) may have reached a saturation point, limiting further effects^23^. Since women have higher SHBG levels than men^24^, it is important to understand how elevated SHBG affects insulin dynamics and T2D risk in WLWH. This is especially relevant for African women who present with a phenotype of low insulin sensitivity and hyperinsulinaemia ^25^.

We hypothesised that higher SHBG and lower total and free testosterone in SA WLWH before and after the menopause will be associated with favourable insulin dynamics that may protect against the development of T2D. Accordingly, the aim of the study was to investigate associations between androgen hormones, SHBG and T2D risk in pre-and post-menopausal Black SA women living with and without HIV.

## Methods

### Design, study population and setting

Recruitment and data collection were conducted between January 2017 and August 2018 at the South African Medical Research Council/Wits Developmental Pathways for Health Research Unit at the Chris Hani Baragwanath Hospital in Soweto, Johannesburg, South Africa. The starting sample for this cross-sectional study included women (n=501) selected from the Middle-Aged Soweto Cohort (MASC)^26^. Exclusion criteria included using menopausal hormone therapy (n=7), hormonal contraceptives (n=30), had a hysterectomy (n=47), had no HIV data (n=1) or blood samples (n=1), were perimenopausal (n=54), resulting in a final sample of 361 participants. The final sample included 81 premenopausal women (57 WLWOH, 24 WLWH), and 280 postmenopausal women (236 WLWOH, 44 WLWH).

The study, conducted in accordance with the declaration of Helsinki, was approved by the Human Research Ethics Committee (HREC) (Medical) of the University of the Witwatersrand (clearance certificate No. M160604). Informed written consent was obtained from all participants.

### Testing procedures

#### Administered questionnaires

Interviewer-administered questionnaires were captured onto REDCap (Version 14.6.7, Vanderbilt University, 2024). Data collected included age, housing density (number of people per room) and asset index (percentage from a possible 12 assets), current medication use, smoking status (current smoker/non-smoker) and alcohol consumption (currently consumes/does not consume). Menopausal status was classified using the date of final menstrual period. Pre-menopause was defined as currently having a regular menstrual cycle and post-menopause was defined as cessation of the menstrual cycle for ≥12 months^27^.

#### HIV testing and CD4 count

Women without a previous HIV positive diagnosis completed a HIV antibody test (One Step HIV–1/2, Guangzhou Wondfo Biotech, China). In WLWH, venous blood was analysed for CD4 count using flow cytometry (Beckman Coulter, Berea, CA, USA). The number of years since HIV diagnosis, number of years on HIV treatment and the medication used were recorded.

#### Body composition

Weight and height were measured using standard techniques. Waist circumference was measured in the mid–axillary line at the midpoint between the lower margin of the last palpable rib and the top of the iliac crest at the end of normal expiration, and hip circumference was measured as the greatest protrusion of the buttocks^28^. Subtotal fat mass (FM) and regional fat distribution were measured using dual energy x-ray absorptiometry (DXA; Hologic Discovery-A (S/N 86254), Bedford, MA, USA, APEX software version 13.4.2:3). Regional fat distribution included trunk and leg FM reported relative (%) to FM^29^. Abdominal visceral (VAT) and subcutaneous adipose tissue (SAT) were estimated^30^. Fat mass index (FMI) was calculated as FM (kg) divided by height squared (m^2^).

#### Fasting venous blood and oral glucose tolerance test (OGTT)

At ∼0800 h and following an overnight fast (10–12 h), baseline venous blood was drawn for the determination of HbA1c, glucose, insulin, C-peptide, SHBG, follicle stimulating hormone (FSH), and luteinizing hormone (LH), E2 and testosterone concentrations. Participants then completed a standard 120-minute OGTT. After ingestion of glucose (75 g), venous blood samples (∼5 mL) were drawn at 30, 60, 90 and 120 minutes for the determination of glucose, and insulin, and C-peptide concentrations.

Participants with known diabetes (on medication) did not complete the OGTT. Homeostasis model assessment of insulin resistance (HOMA-IR) was calculated^31^. Basal insulin clearance was estimated using the ratio of fasting C-peptide to insulin^32^ and peripheral insulin sensitivity was estimated using the Matsuda Index^33^. Insulin response and secretion were estimated using the insulinogenic index (IGI, (ΔInsulin /ΔGlucose))^34^ and C-peptide index (ΔC-peptide /ΔGlucose)^35^, respectively.

Disposition index (DI), which is an estimate of beta-cell function, was calculated (IGI/Matsuda index)^34^. World Health Organization (WHO) criteria were for the classification of glucose tolerance status^36^. The participants were classified as normal glucose tolerance (NGT), impaired glucose metabolism (IGM) including those with impaired fasting glucose (IFG) and/or impaired glucose tolerance (IGT), or T2D^36^. Dysglycaemia was defined as the combination of IGM and T2D.

#### Biochemical analysis

Plasma glucose was analysed on the Randox RX Daytona Chemistry Analyzer using enzymatic methods (Randox Laboratories Ltd., London, UK). Serum insulin and C-peptide was analysed on the Immulite® 1000 Immunoassay System (Siemens Chemiluminescent Healthcare GmbH, Henkestr, Germany). HbA1c levels were measured on whole blood samples using the D-10™ Hemoglobin Analyzer (Bio-Rad Laboratories, Inc., CA, USA). Serum follicle stimulating hormone (FSH), luteinizing hormone (LH) and SHBG were analysed using chemiluminescent microparticle immunoassays (Architect assays, Abbott Laboratories, IL, USA) and albumin was analysed using colorimetric (Bromcresol Green) assay (Alinity c, Abbott Laboratories, IL, USA). Endogenous steroid hormones (E2 and total testosterone) were quantified by ultra-high performance liquid chromatography tandem mass spectrometry (LCMS) ^37, 38^. Free testosterone was calculated ^39^.

### Statistical Analysis

Statistical analysis was performed using STATA version 18 (StatCorp, College Station, Texas). The Shapiro–Wilk test was used to assess the distribution of continuous variables. Unadjusted meanl1±l1standard deviation (SD), median and (25^th^ to 75^th^ percentile), or count (%) are presented for normally distributed, skewed or categorical data, respectively. Within menopausal groups, differences between WLWOH and WLWH were assessed using unpaired t-tests, two-sample Wilcoxon rank sum test or Fisher’s exact test for normally distributed, skewed and categorical data, respectively. Logistic (or multinomial logistic), linear or quantile regressions were used to explore the overall effects of HIV and menopause in the combined sample, including an HIV x menopausal interaction term. Overall group differences in sex hormones as well as glycaemia and insulin parameters were adjusted for age.

Associations between androgens and SHBG, and prevalent dysglycaemia were explored using logistic regression, adjusting for age and FMI. Data reported as odds ratio (OR) (95% confidence interval (CI). In women living without T2D associations between SHBG, and total and free testosterone with continuous glycaemic and insulin parameters, were explored using quantile regression at the 50^th^ percentile, adjusting for age and FMI. Data reported as beta coefficient (β) (95% CI). In all logistic and quantile regressions we explored whether HIV or menopausal status altered these relationships by including interaction terms with sex hormones in the models separately. Age was included in all models to explore the effect of menopausal status independent of chronological age. For both the logistic and quantile regression models, robust Z-scores, calculated from the median and scaled median absolute deviation (MAD), were derived for SHBG, total and free testosterone and the glycaemic and insulin measures to facilitate comparisons of the magnitude of the associations using a standardised measure. The robust Z-scores represent the number of MADs that a data point lies from the median. The MADs were scaled by a constant factor of 1.4826 so that the robust Z-scores are more directly comparable to a traditional Z-score, with 1 robust Z-score indicating a value ∼1 SD from the median. As missing data was minimal (<10%) and missing at random (no differences between groups), pairwise deletion was used when handling missing data.

## Results

### Participant characteristics

Characteristics of participants stratified by HIV and menopausal status are described in Table 1. Postmenopausal women were older than premenopausal women, and within the postmenopausal group, WLWH were younger than WLWOH. Socioeconomic status, characterised by housing density and asset index, did not differ by HIV or menopausal group. Postmenopausal women were less likely to drink alcohol and smoke cigarettes than premenopausal women. Dietary intake and physical activity did not differ between HIV and menopausal groups (data not shown). A greater proportion of postmenopausal women reported taking anti-hypertensive medication compared to premenopausal women, while diabetes and lipid lowering medications did not differ by menopause or HIV. In WLWH, 81% of women were taking antiretroviral therapy (ART), with the majority (73%) using non-nucleoside reverse transcriptase inhibitors (NNRTIs) and this, together with CD4 count, did not differ by menopausal group.

**Table 1.** Participant characteristics.

|  | Premenopausal |  |  | Postmenopausal |  |  | P Value |  |  |
| --- | --- | --- | --- | --- | --- | --- | --- | --- | --- |
|  | WLWOH (n=57) | WLWH (n=24) | P value | WLWOH (n=236) | WLWH (n=44) | P Value | HIV | Meno | HIV x Meno |
| ***Sociodemographic and lifestyle factors*** |  |  |  |  |  |  |  |  |  |
| Age (years) | 48 (46-51) | 46 (45-49) | 0.065 | 58 (54-61) | 54 (50-59) | <0.001 | 0.208 | <0.001 | 0.484 |
| Housing density (people per room) | 1.2 (0.9-1.5) | 1.0 (0.8-1.3) | 0.271 | 1.2 (0.8-1.6) | 1.1 (0.6-1.6) | 0.447 | 0.277 | 0.767 | 0.368 |
| Asset index (% of 12) | 75.0 (66.7-83.3) | 75.0 (58.3-91.6) | 0.664 | 75.0 (58.3-83.3) | 75.0 (58.3-83.3) | 0.871 | >0.999 | >0.999 | >0.999 |
| Current smoker, n (%) | 7 (12.3) | 0 | 0.079 | 10 (4.2) | 2 (4.5) | 0.931 | 0.931 | **0.026** |  |
| Current alcohol consumer, n(%) | 24 (42.1) | 13 (54.2) | 0.320 | 53 (22.5) | 11 (25.0) | 0.712 | 0.321 | **0.003** | 0.578 |
| ***Medication use, n(%)*** |  |  |  |  |  |  |  |  |  |
| Diabetes medications | 4 (7.0) | 1 (4.2) |  | 25 (10.6) | 6 (13.6) |  | 0.630 | 0.421 | 0.501 |
| Hypertension medications | 10 (17.5) | 5 (21.7) |  | 99 (41.9) | 22 (50.0) |  | 0.664 | **0.001** | 0.942 |
| Lipid medications | 3 (5.3) | 1 (4.4) |  | 21 (8.9) | 2 (4.5) |  | 0.865 | 0.371 | 0.710 |
| ***HIV information*** |  |  |  |  |  |  |  |  |  |
| Established on ART, n (%) |  | 18 (75.0) |  |  | 37 (84.1) |  |  | 0.221 |  |
| PIs, n (%) |  | 0 |  |  | 1 (2.3) |  |  | 0.466 |  |
| NNRTIs, n (%) |  | 13 (72.2) |  |  | 27 (73.0) |  |  | 0.721 |  |
| CD4 count (cell/mm<sup>3</sup>) |  | 474 (361-619) |  |  | 611 (416-858) |  |  | 0.128 |  |
| CD4 WCC (cell/mm<sup>3</sup>) |  | 5 (4-6) |  |  | 5 (4-7) |  |  | 0.862 |  |
| CD4 lymphocytes (cell/mm<sup>3</sup>) |  | 33 (21-40) |  |  | 34 (23-40) |  |  | 0.463 |  |
| ***Body composition*** |  |  |  |  |  |  |  |  |  |
| Height (cm) | 158.1 (154.1-162.5) | 157.1 (153.0-160.2) | 0.267 | 158.2 (154.4-162.0) | 156.3 (153.3-160.4) | 0.108 | 0.506 | 0.915 | 0.844 |
| Weight (kg) | 81.8 (77.0-97.4) | 75.6 (63.7-88.3) | 0.065 | 81.3 (72.4-95.1) | 79.9 (64.3-92.8) | 0.106 | 0.755 | 0.873 | 0.846 |
| BMI (kg/m<sup>2</sup>) | 33.3 (29.4-37.7) | 31.6 (26.8-35.7) | 0.121 | 33.1 (29.1-37.8) | 33.1 (26.6-36.5) | 0.184 | 0.173 | 0.868 | 0.271 |
| Waist circumference (cm) | 95.5 (89.6-103.0) | 95.5 (77.5-106.0) | 0.432 | 96.5 (89.6-105.6) | 96.6 (85.1-103.8) | 0.384 | 0.806 | 0.686 | 0.973 |
| Subtotal body fat mass (%) | 43.8 (41.8-46.7) | 43.8 (39.0-45.6) | 0.364 | 46.2 (42.2-49.0) | 44.3 (38.6-49.3) | 0.170 | 0.990 | **0.014** | 0.335 |
| FMI (kg/m<sup>2</sup>) | 14.2±3.5 | 12.5±3.7 | 0.059 | 14.5±4.3 | 13.5±4.8 | 0.151 | 0.102 | 0.683 | 0.570 |
| Leg fat mass (%) | 44.7±7.1 | 43.9±7.2 | 0.681 | 44.2±6.4 | 44.1±7.2 | 0.972 | 0.803 | 0.775 | 0.959 |
| Trunk fat mass (%) | 42.7±6.3 | 42.8±6.1 | 0.924 | 43.2±6.0 | 43.2±6.3 | 0.973 | 0.869 | 0.997 | 0.703 |
| VAT (cm<sup>2</sup>) | 102 (60-128) | 77 (59-108) | 0.223 | 108 (78-140) | 109 (64-132) | 0.385 | 0.128 | 0.518 | 0.196 |
| SAT (cm<sup>2</sup>) | 464 (372-515) | 413 (309-535) | 0.274 | 483 (392-582) | 443 (296-580) | 0.158 | 0.222 | 0.529 | 0.723 |
| VAT/SAT | 0.21 (0.16-0.26) | 0.20 (0.15-0.25) | 0.628 | 0.22 (0.18-0.27) | 0.22 (0.18-0.30) | 0.814 | 0.912 | 0.277 | 0.849 |

Values are median (interquartile range) or mean ± standard deviation. WLWOH, women living without HIV; WLWH, women living with HIV; PI, protease inhibitors, NNRTIs, non-nucleoside reverse transcriptase inhibitors; WCC, white cell count. BMI, body mass index, WHR, waist-hip-ratio; FFSTM, fat-free soft tissue mass; FMI, fat mass index; VAT, visceral adipose tissue; SAT, subcutaneous adipose tissue.

### Differences in body composition

Total body fatness (BMI or body fat %) and body fat distribution (waist circumference, trunk and leg fat mass, VAT and SAT areas) between the menopausal and HIV groups are reported in Table 1. The only difference between the groups was body fat percentage which was higher in postmenopausal compared to premenopausal women. When excluding participants with T2D, weight and FMI were significantly lower in WLWH than WLWOH (p=0.037 and 0.028, respectively, data not shown).

### Differences in SHBG and sex hormones

In age-adjusted analyses LH and FSH were higher and E2 was lower in postmenopausal compared to premenopausal women (Table 2). While LH and FSH did not differ by HIV status, E2 was lower in postmenopausal WLWH than WLWOH (HIV x menopause interaction; p<0.001), while SHBG was higher in postmenopausal WLWH than WLWOH. In contrast, total and free testosterone were lower in pre- and postmenopausal women WLWH compared to WLWOH. Total testosterone was lower in postmenopausal compared to premenopausal women, but free testosterone and SHBG did not differ by menopausal status. The findings did not differ when excluding participants with T2D.

**Table 2.** Differences in SHBG, sex hormones and fasting and OGTT-derived parameters between menopausal and HIV groups.

|  | Premenopausal |  |  | Postmenopausal |  |  | P Value adjusted for age |  |  |
| --- | --- | --- | --- | --- | --- | --- | --- | --- | --- |
|  | WLWOH (n=52) | WLWH (n=21) | P value | WLWOH (n=225) | WLWH (n=41) | P value | HIV | Meno | HIV x Meno |
| **Sex hormones** |  |  |  |  |  |  |  |  |  |
| LH (IU/L) | 6.7 (4.0-14.5) | 4.5 (3.1-12.0) | 0.275 | 21.8 (16.0-29.3) | 22.7 (17.3-31.1) | 0.218 | 0.495 | <0.001 | 0.448 |
| FSH (IU/L) | 11.9 (6.0-23.6) | 8.3 (4.7-18.9) | 0.212 | 55.3 (40.8-69.3) | 63.8 (43.8-75.3) | 0.092 | 0.380 | <0.001 | 0.128 |
| E2 (pmol/L) | 198.0 (77.5-560.5) | 333.0 (96.0-483.0) | 0.991 | **24.0 (15.0-39.0)** | **8.0 (4.3-19.5)** | <0.001 | <0.001 | <0.001 | <0.001 |
| SHBG (nmol/L) | 55.4 (43.4-81.7) | 64.2 (51.3-111.3) | 0.176 | **50.6 (37.8-65.2)** | **72.1 (54.2-103.0)** | <0.001 | 0.326 | 0.370 | 0.180 |
| Total testosterone (nmol/L) | **0.45 (0.25-0.56)** | **0.24 (0.18-0.34)** | **0.008** | **0.35 (0.21-0.57)** | **0.25 (0.09-0.48)** | **0.023** | **0.026** | **0.038** | 0.294 |
| Free testosterone (pmol/L) | **5.4 (3.5-6.9)** | **2.5 (1.3-4.3)** | **0.012** | **4.8 (2.5-8.0)** | **2.0 (1.0-3.6)** | <0.001 | **0.011** | 0.907 | 0.921 |
| **Glycaemic status, n (%)** |  |  |  |  |  |  |  |  |  |
| NGT | 46 (80.7) | 19 (86.4) | 0.703 | 154 (65.8) | 34 (77.3) | 0.065 | Ref | Ref | Ref |
| IGM | 5 (8.8) | 2 (9.1) |  | 44 (18.8) | 2 (4.5) |  | 0.870 | 0.639 | 0.188 |
| T2D | 6 (10.5) | 1 (4.5) |  | 36 (15.4) | 8 (18.2) |  | 0.486 | 0.764 | 0.449 |
| Dysglycaemia | 11 (19.3) | 3 (13.6) | 0.555 | 80 (34.2) | 10 (22.7) | 0.136 | 0.711 | 0.603 | 0.861 |
| **Fasting parameters including participants with T2D** |  |  |  |  |  |  |  |  |  |
| n | 57 | 22 |  | 232 | 44 |  |  |  |  |
| HbA1c (%A1c) | 5.7 (5.3-6.3) | 5.7 (5.3-5.9) | 0.569 | 6.0 (5.6-6.6) | 6.1 (5.7-6.6) | 0.543 | 0.698 | 0.529 | 0.326 |
| Fasting glucose (mmol/L) | 5.0 (4.5-5.5) | 5.0 (4.7-5.5) | 0.577 | 5.0 (4.5-5.7) | 5.2 (4.5-5.8) | 0.592 | 0.753 | 0.262 | 0.604 |
| Fasting insulin (mmol/L) | 8.5 (5.2-14.0) | 10.3 (6.3-14.0) | 0.697 | 9.0 (4.9-14.4) | 9.9 (5.6-16.2) | 0.497 | 0.392 | 0.627 | 0.756 |
| Fasting C-peptide (nmol/L) | 1.6 (1.3-2.3) | 1.8 (1.4-2.2) | 0.710 | 1.8 (1.3-2.5) | 1.9 (1.3-2.7) | 0.457 | 0.512 | 0.522 | 0.828 |
| HOMA-IR | 1.7 (1.2-3.6) | 2.3 (1.3-3.4) | 0.706 | 2.0 (1.1-3.6) | 2.4 (1.3-4.2) | 0.430 | 0.257 | 0.843 | 0.706 |
| Basal insulin clearance | 0.20 (0.15-0.26) | 0.18 (0.16-0.27) | 0.969 | 0.20 (0.16-0.27) | 0.20 (0.15-0.27) | 0.602 | 0.297 | 0.599 | 0.446 |
| **Fasting and OGTT-derived parameters excluding participants with T2D** |  |  |  |  |  |  |  |  |  |
| n | 51 | 22 |  | 199 | 37 |  |  |  |  |
| HbA1c (%A1c) | 5.6 (5.2-6.1) | 5.6 (5.3-5.8) | 0.775 | 6.0 (5.6-6.3) | 6.1 (5.6-6.4) | 0.635 | 0.990 | 0.174 | 0.539 |
| Fasting glucose (mmol/L) | 4.8 (4.4-5.3) | 5.0 (4.7-5.4) | 0.577 | 4.9 (4.4-5.4) | 5.1 (4.5-5.6) | 0.492 | 0.481 | 0.485 | 0.638 |
| Fasting insulin (mmol/L) | 8.0 (5.1-13.8) | 10.2 (6.3-13.4) | 0.623 | 8.1 (4.7-13.2) | 9.6 (5.4-15.7) | 0.357 | 0.301 | 0.518 | 0.814 |
| Fasting C-peptide (nmol/L) | 1.6 (1.3-2.3) | 1.8 (1.4-2.1) | 0.696 | 1.7 (1.3-2.3) | 1.7 (1.3-2.7) | 0.667 | 0.318 | 0.523 | 0.485 |
| HOMA-IR | 1.7 (1.1-3.0) | 2.3 (1.3-2.8) | 0.537 | 1.7 (1.0-3.2) | 2.2 (1.2-3.4) | 0.354 | 0.209 | 0.660 | 0.894 |
| Basal insulin clearance | 0.21 (0.15-0.27) | 0.18 (0.16-0.27) | 0.840 | 0.21 (0.16-0.28) | 0.21 (0.16-0.26) | 0.390 | 0.477 | 0.533 | 0.696 |
| Matsuda index | 5.7 (3.1-8.1) | 4.4 (3.0-8.4) | 0.763 | 5.3 (3.2-8.1) | 4.9 (2.8-8.0) | 0.869 | 0.390 | 0.462 | 0.435 |
| Insulinogenic index | 32.7 (17.5-56.2) | 39.8 (19.6-56.5) | 0.824 | **23.4 (13.0-42.3)** | **38.4 (19.5-65.5)** | **0.002** | 0.582 | 0.490 | 0.357 |
| C-peptide index | 3.2 (1.8-5.6) | 3.6 (2.5-4.3) | 1.000 | **2.8 (1.6-4.4)** | **4.0 (2.4-6.6)** | **0.019** | 0.444 | 0.777 | 0.360 |
| Disposition index | 169.0 (73.5-253.2) | 139.7 (63.3-232.5) | 0.760 | **126.2 (57.1-215.4)** | **214.2 (90.6-461.9)** | **0.007** | 0.418 | 0.622 | **0.019** |

Values are median (interquartile range). WLWOH, women living without HIV; WLWH, women living with HIV; NGT, normal glucose tolerance (fasting plasma glucose <6.1 mmol/L & 120 minute post glucose load <7.8 mmol/L); IFG, impaired fasting glucose (fasting plasma glucose: 6.1–6.9 mmol/L); IGT, impaired glucose tolerance (120 minute post-glucose load: 7.8–11.0 mmol/L); T2D, type 2 diabetes (fasting plasma glucose ≥ 7.0 mmol/L and/or 120 minute post-glucose load ≥ 11.1 mmol/L and/or on diabetes medication. IGM, impaired glucose metabolism (IFG and/or IGT); Dysglycaemia: IGM and/or type 2 diabetes. $S_i$ , insulin sensitivity; $AI R_g$ , acute insulin response to glucose; DI, disposition index; $S_g$ , glucose effectiveness.

### Differences in glycaemic and insulin parameters

Overall, 70.7% of participants presented with NGT, 14.8% with IGM and 14.5% with T2D, and did not differ by HIV or menopausal status (Table 2). The overall prevalence of dysglycaemia was 29.2 (95%CI: 24.6-34.3)%, with 32% of WLWOH presenting with dysglycaemia compared to 20% of WLWH.

Differences in fasting parameters were only evident when excluding those with T2D (n=309). In postmenopausal women without T2D, the IGI, C-peptide index and DI were higher in WLWH compared to WLWOH.

### The association between SHBG, and total and free testosterone with glycaemic and insulin parameters

In the total sample each ∼1SD increase in SHBG was associated with 40% lower odds of dysglycaemia (Table 3), with the association being significant in WLWOH (OR (95% CI): 0.50 (0.35–0.71), p<0.001) but not in WLWH (OR (95%CI): 0.90 (0.62–1.31), p=0.592)(SHBG x HIV interaction p=0.055; Figure 1). The association between SHBG and dysglycaemia did not differ by menopausal status.

**Figure 1.**
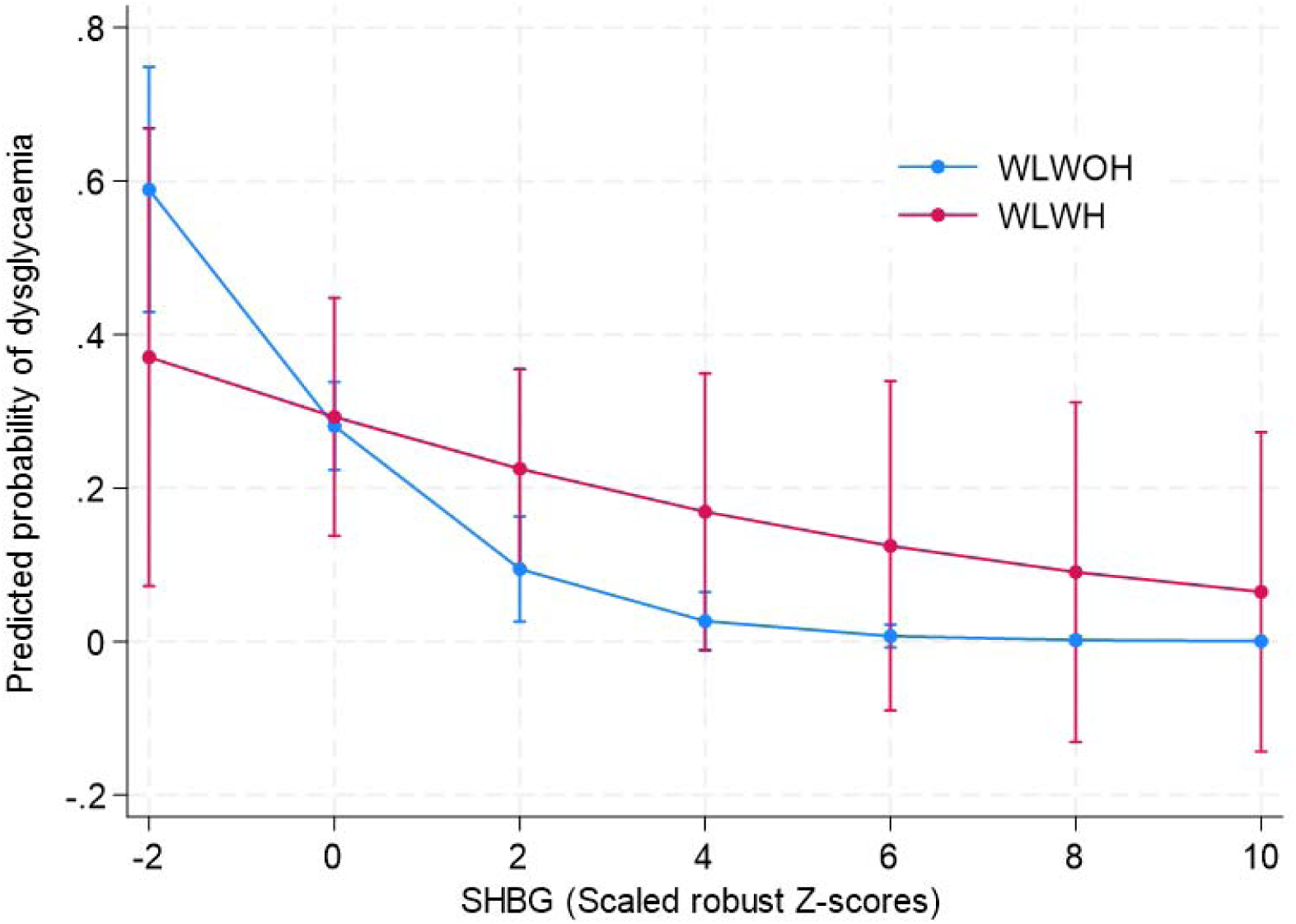
The association between SHBG and prevalent dysglaemia by HIV status (P=0.055 for SHBG x HIV interaction). Higher SHBG was associated with lower odds of dysglycaemia in WLWOH (OR (95% confidence interval (CI): 0.50 (0.35 – 0.71), P<0.001) but not in WLWH (OR (95%CI): 0.90 (0.62 – 1.31, P=0.592). Model adjusted for age and FMI. SHBG presented as scaled robust Z-scores, calculated from the median and median absolute deviation (MAD), with MAD scaled by a constant factor of 1.4826 so that the robust Z-score is comparable to a traditional Z-score.

**Table 3.**
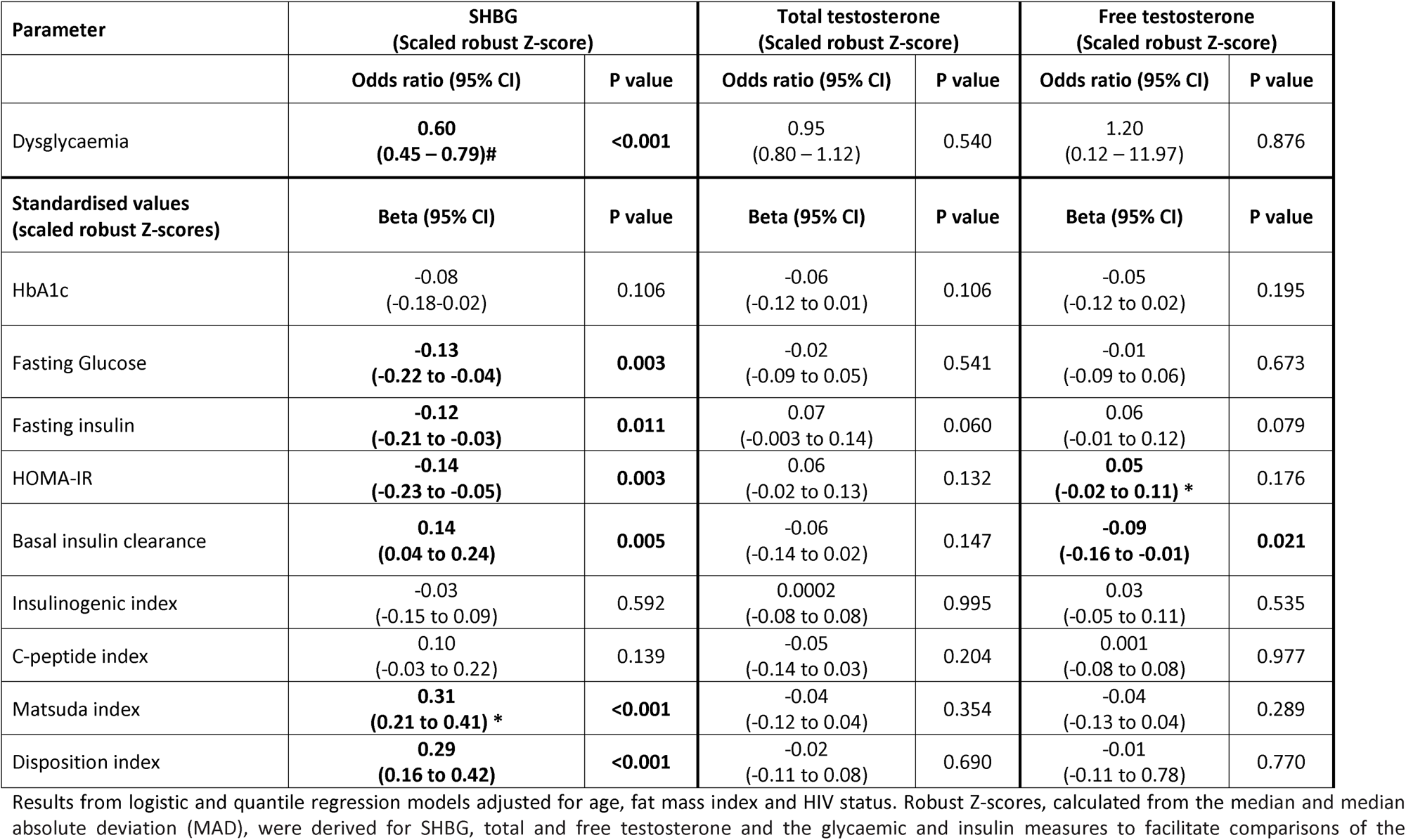
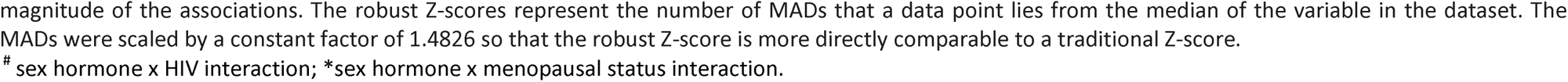
Associations between SHBG, androgen hormones and glycaemic and insulin parameters.

SHBG was positively associated with Matsuda index, DI and basal insulin clearance, and negatively associated with fasting glucose, insulin, and HOMA-IR, irrespective of HIV status, age, and FMI (Table 3). Notably, the association between SHBG and Matsuda index differed by menopausal status (p=0.043 for SHBG x menopause, Figure 2A), with the association stronger in premenopausal (standardised ß (95%CI): 0.46 (0.22-0.70) p<0.001) compared to postmenopausal women (standardised ß (95%CI): 0.29 (0.16-0.43), p<0.001).

**Figure 2.**
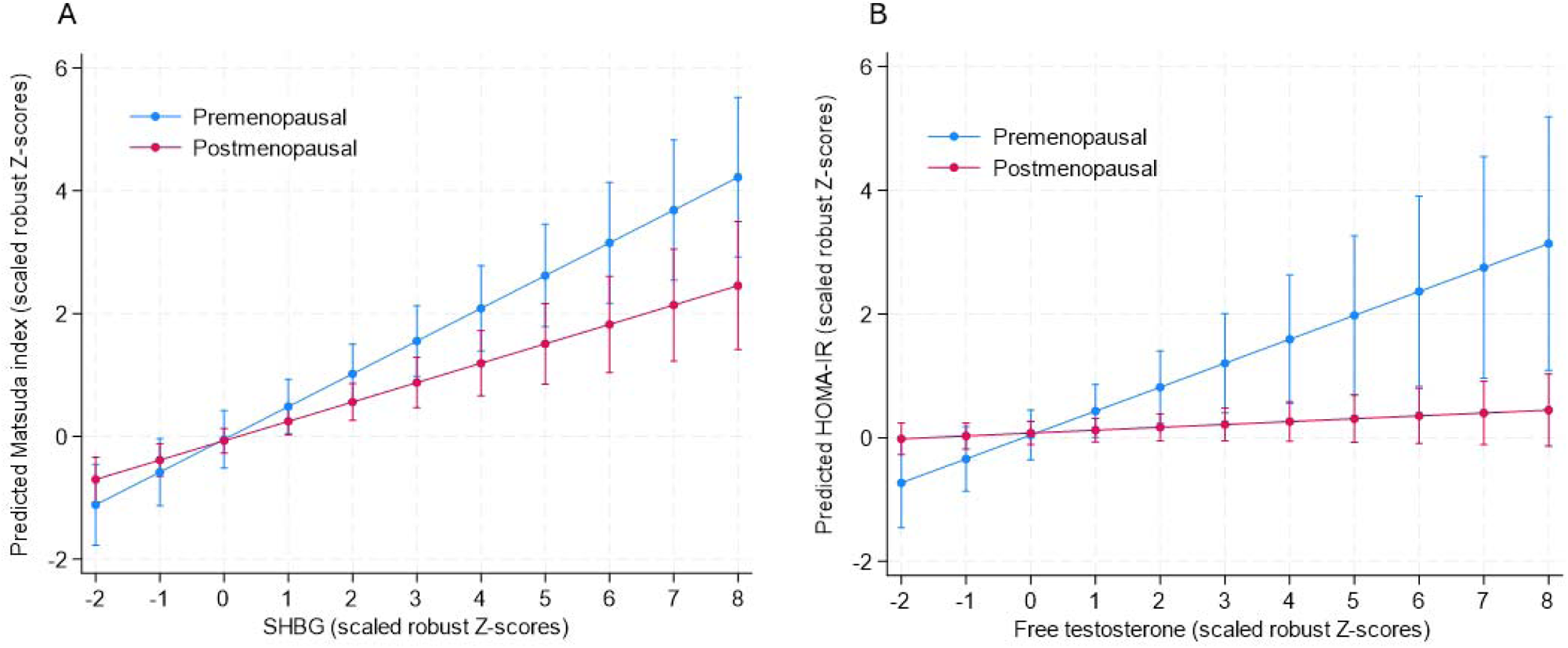
The association between SHBG and free testosterone and insulin parameters by menopausal status, adjusting for age, HIV status and fat mass index. A) The association between SHBG and insulin sensitivity (Matsuda index) differs by menopausal status (p=0.043 for SHBG x menopause), with the association being stronger in premenopausal (standardised ß (95%CI): 0.46 (0.22-0.70) p<0.001) compared to postmenopausal women (standardised ß (95%CI): 0.29 (0.16-0.43), p<0.001). B) The association between free testosterone and HOMA-IR differs by menopausal status (P=0.015 for free testosterone x menopause), with the association only signficiant in the premenopausal women (standardised ß (95%CI): 0.40 (0.13-0.67), p=0.005) and not the post-menopausal women (standardised ß (95%CI):0.05 (-0.02 – 0.12), p=0.200).

Total testosterone was not associated with dysglycaemia or any glycaemic or insulin parameters (Table 3). The association between free testosterone and HOMA-IR was only signficiant in the premenopausal women (standardised ß (95%CI): 0.40 (0.13-0.67), p=0.005) and not the postmenopausal women (standardised ß (95%CI):0.05 (-0.02 – 0.12), p=0.200). Free testosterone was negatively associated with basal insulin clearance, independent of age, HIV status, and FMI.

## Discussion

This is the first study to report sex hormones in Black African women at different stages of the menopausal transition, explore differences by HIV serostatus, and to investigate how SHBG and testosterone levels relate to T2D risk. We showed that SHBG was higher in postmenopausal WLWH compared to WLWOH. Notably, higher SHBG was associated with a T2D-protective phenotype, including higher insulin sensitivity, beta-cell function, and basal insulin clearance, as well as lower fasting glucose, insulin, and HOMA-IR, regardless of HIV status. In contrast, total and free testosterone were lower in both pre- and postmenopausal WLWH than WLWOH. Lower free testosterone was linked to higher basal insulin clearance, and lower HOMA-IR in premenopausal, but not post-menopausal women. These findings support our hypothesis that higher SHBG and lower free testosterone in WLWH compared to WLWOH may be associated with reduced risk of T2D. This study also provides a putative mechanism underlying the lower prevalence of T2D in WLWH and obesity compared to women living with obesity but without HIV^40^. However, longitudinal studies are required to understand the clinical implications of the findings, particularly as premenopausal women transition into post-menopause.

Higher SHBG levels have been consistently and causally associated with lower risk for T2D in both men and women^7, 8, 12, 14–16, 23, 41, 42^. Accordingly, one may hypothesise that the higher SHBG in post-menopausal WLWH may confer reduced risk for T2D compared to WLWOH. Indeed, higher SHBG levels in postmenopausal WLWH were linked to greater beta-cell function compared to WLWOH. Further, SHBG was also associated with lower risk of T2D-related traits, such as lower fasting glucose and insulin, and higher insulin sensitivity, with these associations being independent of fat mass and unaffected by HIV serostatus. Similar alterations in SHBG and androgens have been observed in middle-aged African men, along with significant associations between SHBG, insulin dynamics and incident T2D. However, these associations were only significant in men without HIV and not those living with HIV^23^. The reasons for these differences are unclear, but the association between SHBG and T2D risk is stronger in women, suggesting sexually dimorphic effects^13^. Further, recent results from the KORA study reported higher SHGB levels in women compared to men contributed to sex differences in fasting glucose and incident T2D^24^.

Higher SHBG levels in postmenopausal WLWH than WLWOH is consistent with previous studies^18–20^. Higher SHBG has previously been linked to chronic viral infections including HIV, especially in women and those with HIV RNA >400 copies/mL^20^. The regulation of SHBG is complex, but proinflammatory cytokines like interleukin-1 and tumour necrosis factor alpha reduce SHBG mRNA expression by downregulating its main transcription factor, hepatocyte nuclear factor 4 (reviewed previously^43, 44^). We can only speculate that elevated SHBG levels in WLWH may be a compensatory mechanism to protect against systemic inflammation associated with HIV^20^. SHBG is also regulated by hepatic de novo lipogenesis, adiponectin and sex hormones^43–45^. Mendelian randomisation studies have shown that higher SHBG levels are associated with lower free testosterone, but not total testosterone levels^7^. Indeed, our study shows that higher SHBG in the postmenopausal WLWH was accompanied with lower free testosterone concentrations, which corroborates the findings of WIHS^18^. While E2 is positively associated with SHBG^44^, we showed lower E2 levels in postmenopausal WLWH compared to WLWOH, suggesting that HIV and/or ARVs may directly increase SHBG. Since most of the women were being treated with NNRTI’s it is difficult to differentiate the effect of HIV from those of ARVs on endocrine function. Further research is needed to better understand the mechanisms underlying this endocrine dysregulation in WLWH over the menopausal transition.

Nonetheless, our study showed that the association between SHBG and prevalent dysglycaemia was only significant in WLWOH. We postulate that the lower total and free testosterone concentrations in WLWH may confer reduced risk for T2D, with higher levels of SHBG having no additional benefit. In contrast, in WLWOH who present with higher total and free testosterone levels compared to WLWH, higher SHBG may reduce T2D risk by lowering the bioavailability of testosterone. This is supported by our findings, which showed that the association between SHBG and insulin sensitivity was stronger in premenopausal women, who have higher total testosterone levels compared to postmenopausal women (Figure 2A). A recent Mendelian randomisation study showed a causal link between the bioavailability of testosterone and increased risk of T2D in women^7^. Similar to our study and others^14, 16, 21, 42, 46^, the Mendelian randomisation study showed no association between total testosterone and T2D risk, suggesting that the associations between SHBG and bioavailable testosterone is likely driven by SHBG, either directly or in combination with free testosterone^7^. The exact mechanisms by which SHBG may influence T2D risk are not fully understood. However, SHBG has been shown to act as a hepatokine, mediating the link between intrahepatic lipids, insulin sensitivity and T2D status, with stronger effects in women than men^47, 48^. Further, SHBG binds to GPRC6A, stimulating insulin release in a dose dependent manner^49^, providing a potential mechanism for the observed relationships between SHBG and beta-cell function.

Our study showed that free testosterone was associated with lower basal insulin clearance and higher HOMA-IR, with this association only significant in premenopausal women. This could be due to the lower total androgen load after menopause. These results are supported by other studies showing that higher free testosterone in premenopausal women is associated with reduced hepatic insulin clearance and lower insulin sensitivity, but not with obesity-related insulin secretion^50^. Other studies also suggest that the relationship between free testosterone and incident T2D is mediated by adiposity and insulin resistance^42^. Taken together, our results indicated that higher SHBG, coupled with lower free testosterone levels may confer reduced risk for T2D in WLWH than WLWOH.

Despite significantly lower E2 in the postmenopausal women, no differences in body composition or T2D risk were observed between the menopausal groups, findings contrary to others^51^. However, others have shown that Black African women may not gain as much abdominal adiposity across the menopause transition due to both higher abdominal adiposity and smaller fluctuations in sex steroid hormones in the years leading up to menopause^52^. In our study, the majority (67.1%) of the women were living with obesity, limiting the potential for further increases in body fat and related changes in glycaemic and insulin parameters. A comparative study conducted in SSA revealed no significant differences in anthropometric and cardiometabolic risk factors between pre- and post-menopausal women from South and East Africa who presented with a high prevalence of obesity (32-66%)^5, 53^. In contrast, women from West Africa, with a lower prevalence of obesity (1-5%), showed distinct variations in cardiometabolic risk between menopausal stages^5^.

### Strengths and limitations

This study comprehensively phenotypes Black African women, including OGTT-derived measures of glycaemic and insulin parameters, as well as detailed characterisation of sex hormones measured using LCMS. The study is however limited by the cross-sectional design that precludes inferences about causality. Free testosterone was not measured, and we relied on calculated free testosterone^39^. In addition, the relatively small sample of premenopausal women and WLWH limits our analysis and interpretation of the results. Further, sex hormone measurements in premenopausal women were not taken at a set time during the menstrual cycle, however we do not believe that the phase of menstrual cycle influenced the androgen results as differences in these hormones by HIV serostatus were consistent at each menopausal stage.

## Conclusion

For the first time, we show that midlife Black African WLWH have higher SHBG, and lower total and free testosterone concentrations compared to WLWOH, which may confer reduced risk for T2D. These findings offer valuable insights into the complex relationships between androgenic hormones, HIV, menopause, and T2D risk. However, longitudinal studies are required to understand the clinical implications of these associations.

## Acknowledgements

We are grateful to the participants of the Middle-Aged Soweto Cohort and the research staff who were involved in this study.

## Data availability

Data is available from authors upon reasonable request

