## Supplementary figures and images for "Sex hormone-binding globulin, testosterone and type 2 diabetes risk in middle-aged African women: exploring the impact of HIV and menopause"

### Fig 1

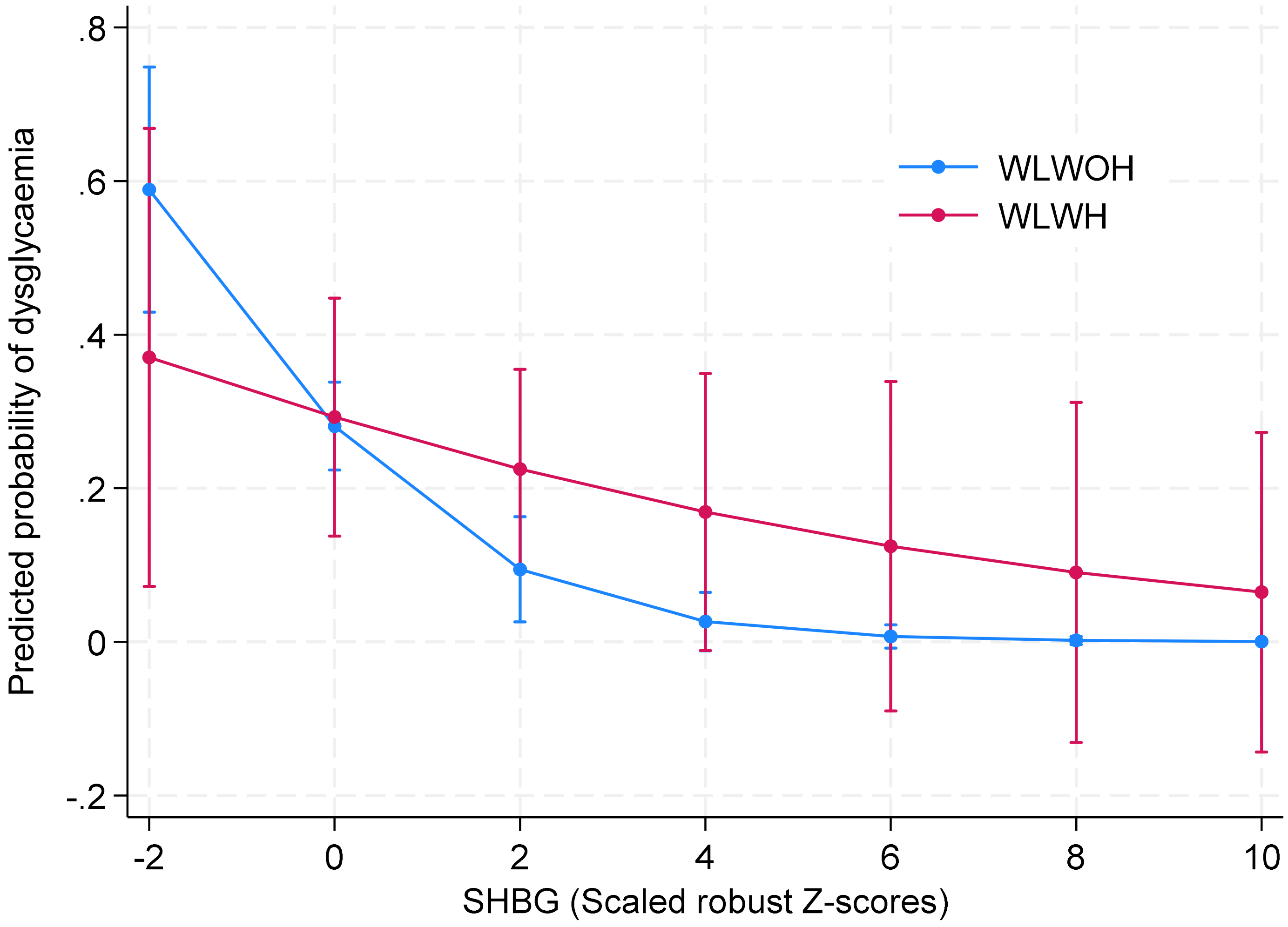

### Fig 2

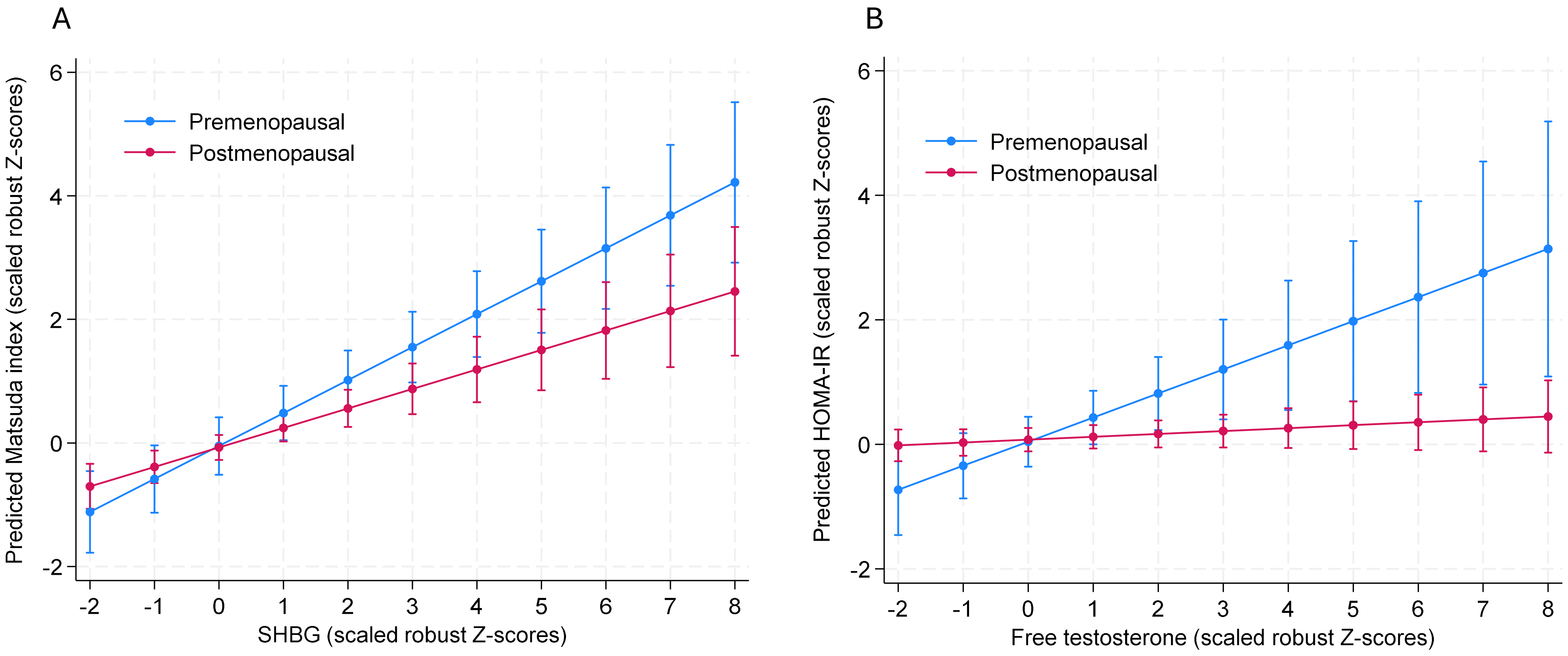
